# COVID-19 Mortality in Cancer Patients: A Report from a Tertiary Cancer Centre in India

**DOI:** 10.1101/2020.09.14.20194092

**Authors:** Anurag Mehta, Smreti Vasudevan, Anuj Parkash, Anurag Sharma, Tanu Vashist, Vidya Krishna

## Abstract

**Background:** Cancer patients, especially those receiving cytotoxic therapy are assumed to have a higher probability of death from COVID-19. We have conducted this study to identify the Case Fatality Rate (CFR) in cancer patients with COVID-19 and have explored the relationship of various clinical factors to mortality in our patient cohort.

**Methods:** All active cancer cases presented to the hospital from 8^th^ June to 24 August 2020, and developed symptoms/ radiological features suspicious of COVID-19 were tested by Real-time polymerase chain reaction assay and/or cartridge-based nucleic acid amplification test from a combination of naso-oropharyngeal swab for SARS-CoV-2. Clinical data, treatment details, and outcomes were assessed from the medical records.

**Results:** Of the total 3101 cancer patients admitted to the hospital, 1088 patients were tested and 186 patients were positive for SARS-CoV-2. The CFR in the cohort was 27/186 (14.5%). Univariate analysis showed that the risk of death was significantly associated with the presence of comorbidities [OR: 2.68; (95%CI: 1.13–6.32); *P* = 0.02], multiple comorbidities [OR: 3.01; (95%CI: 1.02–9.07); *P* = 0.046 for multiple vs. single], and the severity of COVID-19 presentation [OR: 27.48; (95%CI: 5.34–141.49); *P* = 0.0001 for severe vs. not severe]. Among all comorbidities, diabetes [OR: 3.3; (95%CI: 1.35–8.09); *P* = 0.008] and cardiovascular diseases [OR: 3.77; (95%CI: 1.02–13.91); *P* = 0.045] were significant risk factors for death. The receipt of anticancer treatments including chemotherapy, surgery, radiotherapy, targeted therapy, and immunotherapy within a month before the onset of COVID-19 symptoms had no significant effect on the mortality of cancer patients.

**Conclusion:** To the best of our knowledge, this is the first study from India reporting the CFR, clinical associations, and risk factors for mortality in SARS-CoV-2 infected cancer patients. Our study shows that the frequency of COVID-19 in cancer patients is high, and the CFR is 7.6 times more than the national average. Anticancer therapies did not increase the risk of death. Pre-existing comorbidities specially diabetes, multiple comorbidities, and severity of COVID-19 presenting symptoms are significantly linked with COVID-19 related death in the cohort.

## Introduction

The ongoing pandemic of coronavirus disease 2019 (COVID-19) caused by severe acute respiratory syndrome coronavirus 2 (SARS-CoV-2) has caused unprecedented health and societal crises across the globe. This catastrophe however has victimized the cancer patients the most, adversely impacting diagnosis and treatment in about 55% of cases worldwide
^1^
. It is generally assumed that patients with cancer are at a higher risk of contracting COVID-19. Additionally, it is also postulated that the severity and resulting mortality is amplified in cancer patients with COVID-19 due to their elderly and the immunocompromised state further worsened by cancer treatment. Multiple studies from different geographical locations show that the case fatality rate (CFR) of the SARS-CoV-2 infected cancer patients varies from 3.7% to 61.5%^(2–4)^. Despite recent efforts, no clear consensus has been reached for the relation of mortality to demographics, cancer type, stage, and underlying comorbidities; and it may depend on the epidemiology and prevalent oncology practices^(5, 6)^. Various studies regarding the risk of treating cancer patients during the pandemic have shown contradictory results^(7–10)^. Larger datasets acquired globally from different sources and geographic locations and rationally analyzed has the best potential to provide an effective risk-benefit calculus to assist oncologists to optimize the management and use anticancer treatments to restore outcomes to the pre-COVID 19 eras. We aim to describe the clinical and demographic characteristics and COVID-19 outcomes in a cohort of patients with cancer and symptomatic COVID-19. An attempt has also been made to assess the adverse effect of cytotoxic and other novel therapies on mortality in cancer patients with COVID-19, if any.

## Material and Methods

### Study design and subjects

The study is single-center, retrospective, conducted at a tertiary cancer care hospital. Patients with active cancer presented to the hospital between 8^th^ June to 20^th^August 2020, and with confirmed COVID-19 infection were included. Clinical data, anticancer treatment details (performed within a month of COVID-19 diagnosis), clinical course, and outcome were retrieved from the hospital electronic medical records. The COVID-19 infection severity of patients was scored at the time of presentation by the treating physician according to the Ministry of Health and Family Welfare (Government of India) guidelines^(11, 12)^.

The study has been approved by our Institutional Review Board (RGCIRC/IRB-BHR/61/2020) and was conducted according to the Declaration of Helsinki.

### Real Time Polymerase Chain Reaction assay

According to the Indian Council of Medical Research (ICMR) guidelines and international practice, the COVID-19 symptomatic cancer patients were tested by Real-Time Polymerase Chain Reaction (RT-PCR) assay and/or cartridge-based nucleic acid amplification test (CBNAAT) test for SARS-CoV-2.

Samples were collected from the nasopharynx and oropharynx in a single tube with 3 ml of the viral transport medium (Biogenix^®^Lucknow - 226012, Uttar Pradesh, India). The RT-PCR test was performed on ‘QuantStudio™ 5 Real-Time PCR System (Thermo-Fishers Scientific-Life Technologies Holdings Pvt Ltd, Block 33, Marsiling Industrial Estate Road 3, Singapore 739256) employing an ICMR approved RT-PCR test kit (TRUPCR^®^-3B BLACKBIO Biotech, Govindpura, Bhopal, Madhya Pradesh 462023, India). This kit detects E gene common to the Sarbecovirus superfamily, a sampling control of human ribonucleic acid nuclease P (hRNaseP) and RNA dependent RNA polymerase (RdRP) gene, and N gene for the detection of SARS-CoV-2. The test was performed and interpreted as per manufacturer’s instructions.

Rapid molecular testing was used in certain cases utilizing a cartridge-based nucleic acid testing (CBNAAT) from Cepheid^®^USA, Sunnyvale, CA 94089, United States (GeneXpert^®^assay). It identifies E gene-specific RNA and N2 gene-specific RNA. The test was performed and interpreted as per the downloaded package insert.

### Statistical Analysis

Continuous variables were presented as mean±SD or median (interquartile range, IQR). Categorical variables were presented as frequencies and percentages. The two-sided independent t-test and the median test was used to compare the mean and median ages, respectively. Fisher’s exact test/ Pearson’s Chi-Squared test was used to compare categorical data. Univariate logistic regression was used to estimate Odds Ratios (OR) and 95% Confidence Intervals (CI). Multivariate logistic regression was used to compute odds ratio for the various treatment modalities after adjusting for age and comorbidities. All the statistical analyses have performed either by using SPSS^®^ Version 23.0 software or MedCalc Statistical Software version 19.4.0. The reported *P* values are two-sided and a *P* value< 0.05 was considered statistically significant.

## Results

A total of 3101 cancer patients were treated at the indoor facility of the center. One thousand and eighty-eight patients developed signs, symptoms, and/ or radiological features suspicious of COVID-19. Of these 186 tested positive for COVID-19 and formed the study cohort. The infection rate of COVID-19 among all cancer patients treated at the center was ∼6% (186/3101) and 17.1% (186/1088) of the symptomatic and tested cancer patients. The clinical features are shown in Table 1. Most patients had solid malignancies (82.3%); gastrointestinal cancer (21.5%) was the commonest cancer type. About 17.7% of cases presented with hematological malignancies. More than a quarter of cases (26.9%) were metastatic. Eighty-six patients (46.2%) presented at least a single comorbidity; hypertension(24.2%) and diabetes (18.3%) were the commonest. About 60% of cases were on active cancer treatment and had received cancer-directed treatment within a month before the onset of COVID-19 symptoms. The majority of patients were on chemotherapy (37%).

**Table 1.** Clinical characteristics of cancer patients infected with COVID-19 according to outcome (N = 186)

|  | **Total**<br>**N = 186 (%)** | **Survivors**<br>**n = 159 (%)** | **Non-survivors**<br>**n = 27 (%)** | **P value** |
| --- | --- | --- | --- | --- |
| **Age (years)** |  |  |  |  |
| Mean±SD | 50.24±15.77 | 50.22±15.66 | 50.33±16.75 | 0.97 |
| Median (IQR) | 52 (42-58.75) | 52 (42-58.5) | 53 (43.5-60) | 0.95 |
| **Gender** |  |  |  |  |
| Male | 105 (56.4) | 89 (56) | 16 (59.3) | 0.83 |
| Female | 81 (43.6) | 70 (44) | 11 (40.7) |  |
| **Comorbidities** |  |  |  |  |
| Present | 86 (46.2) | 68 (42.8) | 18 (66.7) | 0.035 |
| Absent | 100 (53.8) | 91 (57.2) | 9 (33.3) |  |
| **Number of comorbidities in a patient** |  |  |  |  |
| No comorbidity | 100 (53.8) | 91 (57.2) | 9 (33.3) | 0.004 |
| Single | 47 (25.3) | 41 (25.8) | 6 (22.2) |  |
| More than one | 39 (21.0) | 27 (17.0) | 12 (44.4) |  |
| **Type of comorbidities** |  |  |  |  |
| Cardiovascular disease | 11 (5.9) | 7 (4.4) | 4 (14.8) | 0.06 |
| Chronic obstructive pulmonary disease | 2 (1.1) | 2 (1.3) | 0 (0) | 1 |
| Diabetes | 34 (18.3) | 24 (15.1) | 10 (37.0) | 0.01 |
| Hypertension | 45 (24.2) | 36 (22.6) | 9 (33.3) | 0.23 |
| Thyroid (Hypo/ Hyper) | 30 (16.1) | 26 (16.4) | 4 (14.8) | 1.00 |
| Other comorbidities | 5 (2.7) | 3 (1.89) | 2 (7.1) | 0.15 |
| **Solid vs Hematological cancer** |  |  |  |  |
| Solid | 153 (82.3) | 133 (83.7) | 20 (74.1) | 0.27 |
| Hematological | 33 (17.7) | 26 (16.3) | 7 (25.9) |  |
| **Cancer type** |  |  |  |  |
| Brain | 2 (1.1) | 2 (1.3) | 0 (0) | 0.09 |
| Head and neck | 33 (17.7) | 27 (17) | 6 (22.2) |  |
| Breast | 19 (10.2) | 18 (11.3) | 1 (3.7) |  |
| Thoracic | 17 (9.1) | 17 (10.7) | 0 (0) |  |
| Musculoskeletal and skin | 6 (3.2) | 4 (2.5) | 2 (7.4) |  |
| Gastrointestinal | 40 (21.5) | 31 (19.5) | 9 (33.3) |  |
| Genitourinary and gynecologic | 36 (19.4) | 34 (21.4) | 2 (7.4) |  |
| Hematological | 33 (17.7) | 26 (16.3) | 7 (25.9) |  |
| **Cancer spread** |  |  |  |  |
| Localized tumor | 58 (31.2) | 52 (32.7) | 6 (22.2) | 0.36 |
| Locally advanced | 78 (41.9) | 67 (42.1) | 11 (40.7) |  |
| Metastatic | 50 (26.9) | 40 (25.2) | 10 (37.1) |  |
| **Cancer directed treatment within 1 month of COVID-19 infection** |  |  |  |  |
| Treated cases | 112 (60.2) | 94 (59.1) | 18 (66.7) | 0.53 |
| No treatment | 74 (39.8) | 65 (40.9) | 9 (33.3) |  |
| **Cancer treatment** |  |  |  |  |
| Surgery | 31 (16.7) | 27 (17) | 4 (14.8) | 1.00 |
| Chemotherapy | 69 (37.1) | 56 (35.2) | 13 (48.1) | 0.20 |
| Radiotherapy | 21 (11.3) | 20 (12.6) | 1 (3.7) | 0.32 |
| Targeted therapy | 6 (3.2) | 4 (2.5) | 2 (7.4) | 0.21 |
| Immunotherapy | 11 (5.9) | 10 (6.3) | 1 (3.7) | 1.00 |
| **Severity of COVID-19 infection** |  |  |  |  |
| Mild | 134 (72) | 125 (78.6) | 9 (33.3) | <0.0001 |
| Moderate | 43 (23.1) | 32 (20.1) | 11 (40.7) |  |
| Severe | 9 (4.9) | 2 (1.3) | 7 (26) |  |
| **Patients who received ventilator support** | 12 (6.4) | 0 (0) | 12 (44.4) | 0.00 |

The chief presenting symptoms of SARS-CoV-2 infection were fever (123/186, 66.1%, fatigue (26/186, 14%) and respiratory distress (25/186, 13.4%). The disease was presented severe in 9/186 (∼5%) cases. The COVID-19 associated fatality rate in the cohort was 14.5% (27/ 186) [median follow-up duration: 63 days]. The CFR for hematological malignancies tended to be higher than the CFR for solid malignancies (7/33, 21.2% vs. 20/153, 13.1%).

Severe COVID-19 infected cancer cases were managed by treatments including corticosteroids, hydroxychloroquine, remdesivir, tocilizumab and convalescent plasma therapy. Assisted ventilation had to be given to 12 patients (6.4%), however, all of them eventually developed COVID-19 related complications like pneumonitis and associated respiratory failure, septic shock or sudden cardiac arrest and succumbed to the disease.

Next, we explored the differences between the cancer patients who died and those who survived the SARS-CoV-2 infection (Table 1). There was no significant difference between the survivors and the non-survivors with respect to age, gender, type of malignancy, and cancer spread. Also, no significant effect on mortality was noted for the patients who had received anticancer therapy within the past month. Deceased patients displayed significantly higher rates of comorbidity compared to the cancer patients who survived(66.7% vs. 42.8%, *P* = 0.035); patients with greater than one comorbidity had significantly inferior outcome than those with single or no comorbidity (*P* = 0.004). Importantly, patients with diabetes experienced significantly more deaths than patients without diabetes (*P* = 0.01) (Table 1). Further, we observed that cancer patients presented with moderate and severe COVID-19 symptoms had significantly higher mortality than those presented with mild symptoms (*P*< 0.0001).

The logistic regression analysis (univariate) for death has been shown in Table 2. The odds ratio was statistically significant for the presence of comorbidity [OR = 2.68, *P* = 0.02], multiple vs. single morbidity [OR = 3.01, *P* = 0.046], cardiovascular disease [OR = 3.77, *P* = 0.045] and diabetes [OR = 3.3, *P* = 0.008]. The odds of death were significantly higher in patients presented with severe COVID-19 infection symptoms in comparison to mild/moderately symptomatic patients (OR = 27.48, *P* = 0.0001). Patients who were on active cancer treatment during one month span before contracting COVID-19 did not have an increased risk of death. Also, the treatment modalities including surgery, chemotherapy, radiotherapy, targeted therapy, and immunotherapy did not confer an increased risk of death in univariate analysis. There was a significant difference in the median age of the patients who received chemotherapy compared to those who did not (49 years vs. 54 years, *P* = 0.005). So we further examined whether anticancer therapies could influence mortality in the cohort by adjusting for age and comorbidity in the logistic regression analysis (multivariate) (Supplementary table 1). Compared to the patients who were not on therapy, there was no significant increased risk of death with chemotherapy (OR = 1.63, *P* = 0.30), radiotherapy (OR = 0.19, *P* = 0.13), targeted therapy (OR = 2.70, *P* = 0.29), immunotherapy (OR = 0.40, *P* = 0.41), or surgery (OR = 0.87, *P* = 0.82) (Supplementary table 1).

**Table 2.** Logistic regression analysis (univariate) and odds ratio for death in the cohort (N = 186 COVID-19 infected cancer patients)

| Variable | Odds ratio | 95% CI | P value |
| --- | --- | --- | --- |
| Age | 1 | 0.97 - 1.03 | 0.97 |
| Gender | 1.14 | 0.49 - 2.62 | 0.75 |
| Comorbidity | 2.68 | 1.13 - 6.32 | 0.02 |
| Comorbidity (multiple vs. single) | 3.01 | 1.02 - 9.07 | 0.046 |
| Cardiovascular disease | 3.77 | 1.02 - 13.91 | 0.045 |
| Diabetes | 3.3 | 1.35 - 8.09 | 0.008 |
| Hypertension | 1.71 | 0.71 - 4.13 | 0.23 |
| Thyroid (hypo/ hyper) | 0.89 | 0.28 - 2.79 | 0.84 |
| Other comorbidities | 4.16 | 0.66 - 26.15 | 0.13 |
| Hematological malignancies vs. solid tumors | 1.79 | 0.69 - 4.67 | 0.23 |
| Head and neck | 1.4 | 0.51 - 3.79 | 0.51 |
| Breast | 0.3 | 0.04 - 2.36 | 0.25 |
| Musculoskeletal and skin | 3.1 | 0.54 - 17.82 | 0.2 |
| Gastrointestinal | 2.06 | 0.85 - 5.03 | 0.11 |
| Genitourinary and gynecologic | 0.29 | 0.07 - 1.30 | 0.11 |
| Hematological | 1.79 | 0.68 - 4.67 | 1.79 |
| Advanced vs. localized cancer | 1.42 | 0.50 - 4.10 | 0.51 |
| Metastatic vs. localized cancer | 2.17 | 0.73 - 6.46 | 0.16 |
| Cancer treatment | 1.38 | 0.58 - 3.27 | 0.46 |
| Surgery | 0.85 | 0.27 - 2.66 | 0.78 |
| Chemotherapy | 1.71 | 0.75 - 3.88 | 0.2 |
| Radiotherapy | 0.27 | 0.03 - 2.08 | 0.21 |
| Targeted therapy | 3.1 | 0.54 - 17.82 | 0.20 |
| Immunotherapy | 0.57 | 0.07 - 4.67 | 0.60 |
| Mild COVID-19 infection | 0.14 | 0.056 – 0.33 | <0.0001 |
| Moderate COVID-19 infection | 2.72 | 1.15 – 6.45 | 0.02 |
| Severe COVID-19 infection | 27.48 | 5.34 – 141.49 | 0.0001 |

Male was compared with female. Hematological malignancies were compared with solid tumors. For other categorical variables absence was taken as a reference.

## Discussion

The COVID-19 pandemic has raised several fears. One among these has been the increased propensity of cancer patients to contract COVID-19. In the current study, we found the incidence of COVID-19 to be ∼6.0% against the national incidence of 0.32% in the general population and 0.29% as per European Centre for Disease Prevention and Control (Figure 1a), the incidence in cancer patients is (>)15 fold higher^(13, 14)^. This on one hand can be ascribed to patients’ factors, visitations, and admission to a health care facility with a high risk of contracting infection; may also be partly because of low testing rates in India and poor access to health care facilities. However, even when compared to the reported incidence in other high COVID-19 incidence nations like Brazil and the USA, the incidence is still 3 times higher and several-fold more than middle and low incidence countries (Figure 1b). While the present study does not explore the reasons for this alarmingly high occurrence of COVID-19 in cancer patients, it does establish the higher incidence of COVID-19 in cancer patients.

**Figure 1(a).**
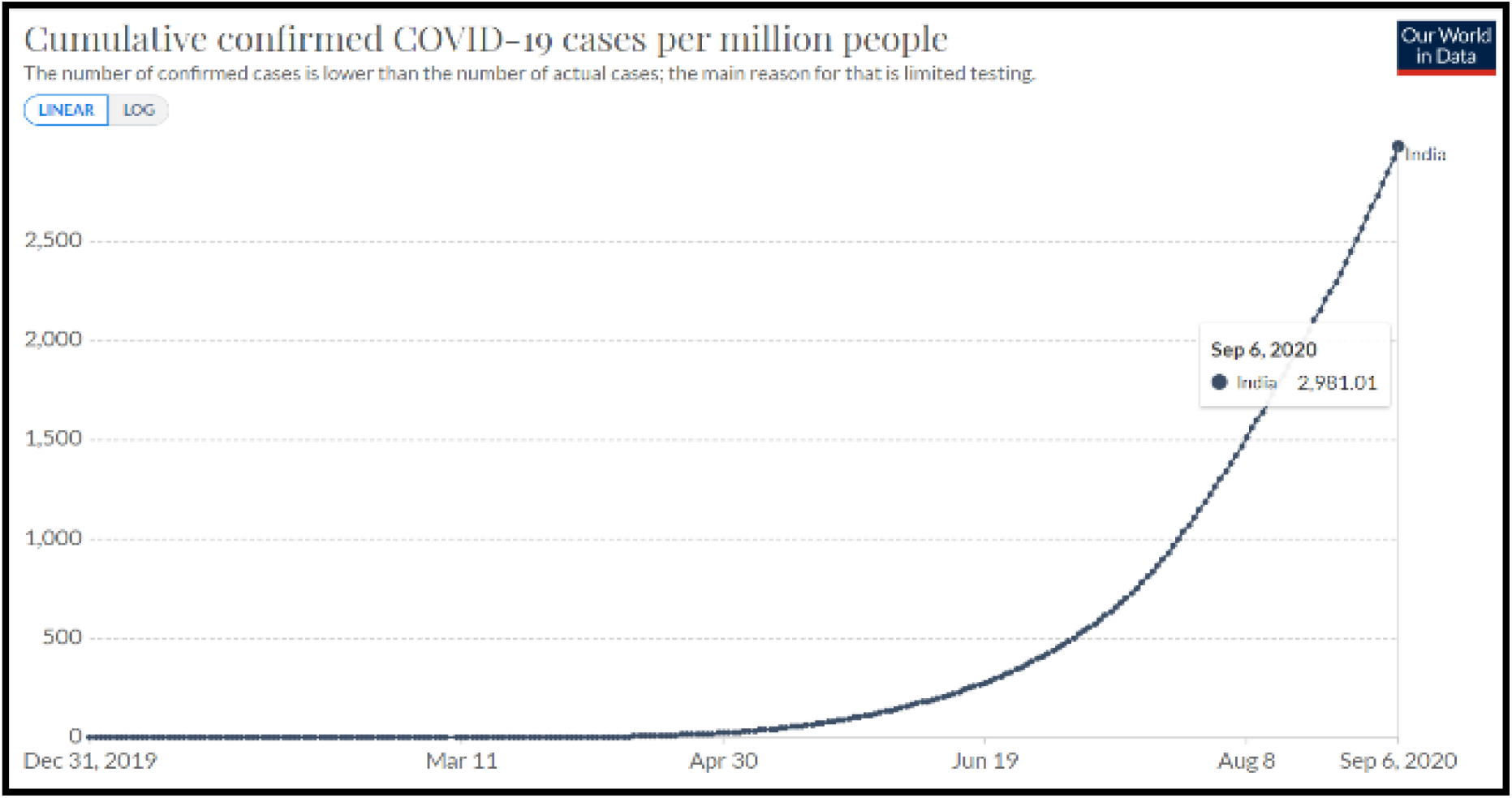
Cumulative COVID-19 incidence per million population (India) Source: European Centre for Disease Prevention and Control (Data as on September 6, 2020)^(14)^

**Figure 1(a).**
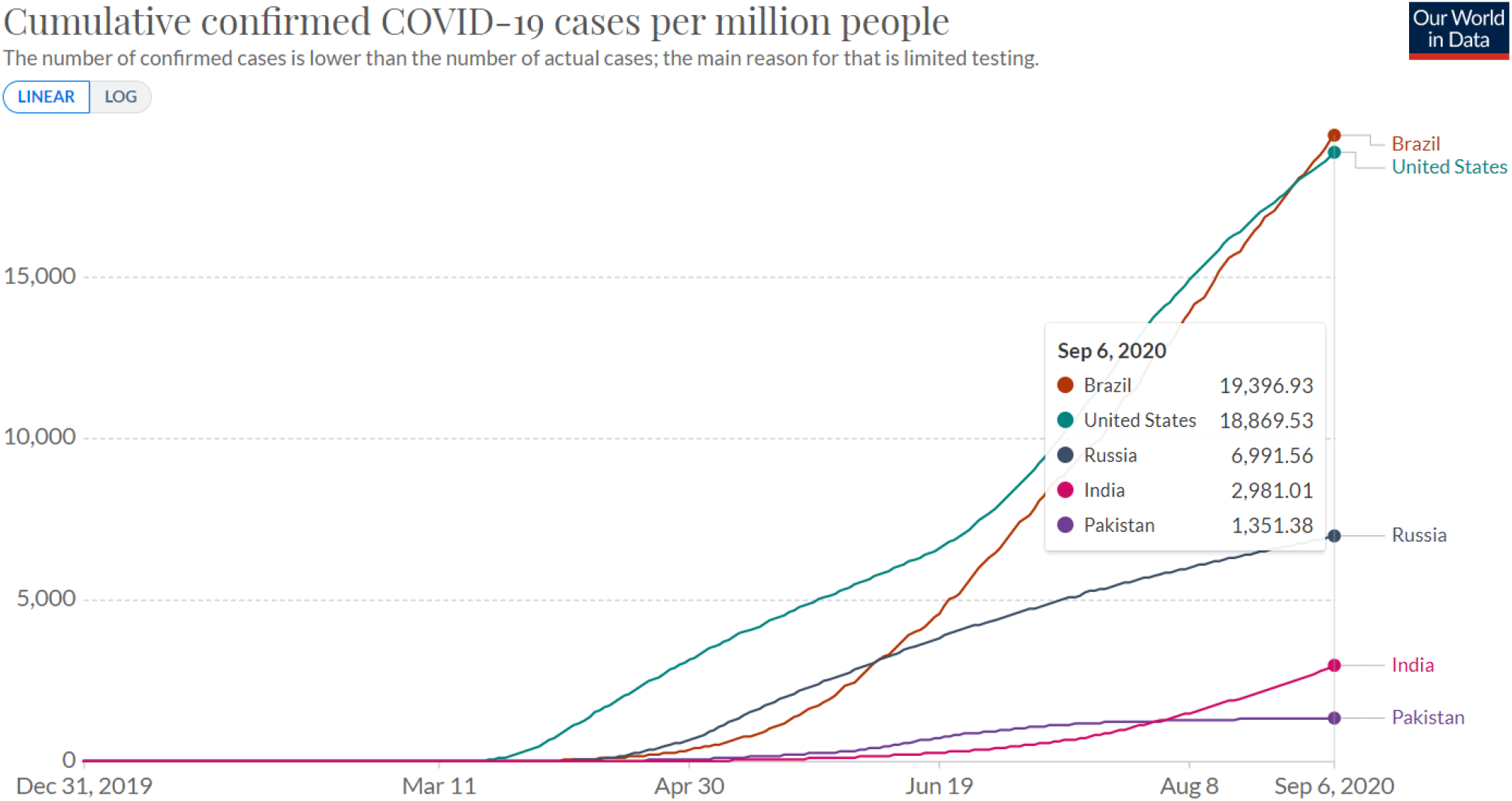
The incidence of COVID 19 in general population ranges from 1.9% in Brazil to 0.13% in Pakistan (Data as on September 6, 2020) Source: European Centre for Disease Prevention and Control (Data as on September 6, 2020)^(14)^

The CFR in the present study was observed to be 14.5 % against the national average of 1.9% in the general population (7.6 times higher) as per the national database (Arogyasetu Application), 1.84% by John Hopkin COVID-19 tracker, and 1.7% as per the European Centre for Disease Prevention and Control (Figure 1c)^(13–15)^. Two recent studies from the European continent have shown a far greater CFR in cancer patients with COVID-19. In the UK Coronavirus Cancer Monitoring Project, a CFR of 30.6% was observed, where 319 of the 1044 cancer patients with COVID-19 died with 92.5% had their death attributed directly to COVID-19^(9)^. In another observational study by Pinato *et al*. of 890 cancer patients diagnosed with SARS-CoV-2, mortality was found to be 33.6%^(10)^. Similarly, high rates were noticed in a New York Hospital System where a CFR of 28% was observed^(16)^. A large systematic review of 52 studies by a group led by Kamal S Saini involving more than 18,000 cases of cancer with COVID-19, the probability of death was 25·6% (95% CI 22·0% to 29·9%; I^2^ = 48·9%)^(4)^. Relatively, lower CFR in the present study when contextualized to CFR in the general population shows the similar proportion to those observed elsewhere. This study once again establishes the far higher CFR in cancer patients with COVID-19.

**Figure 1(c).**
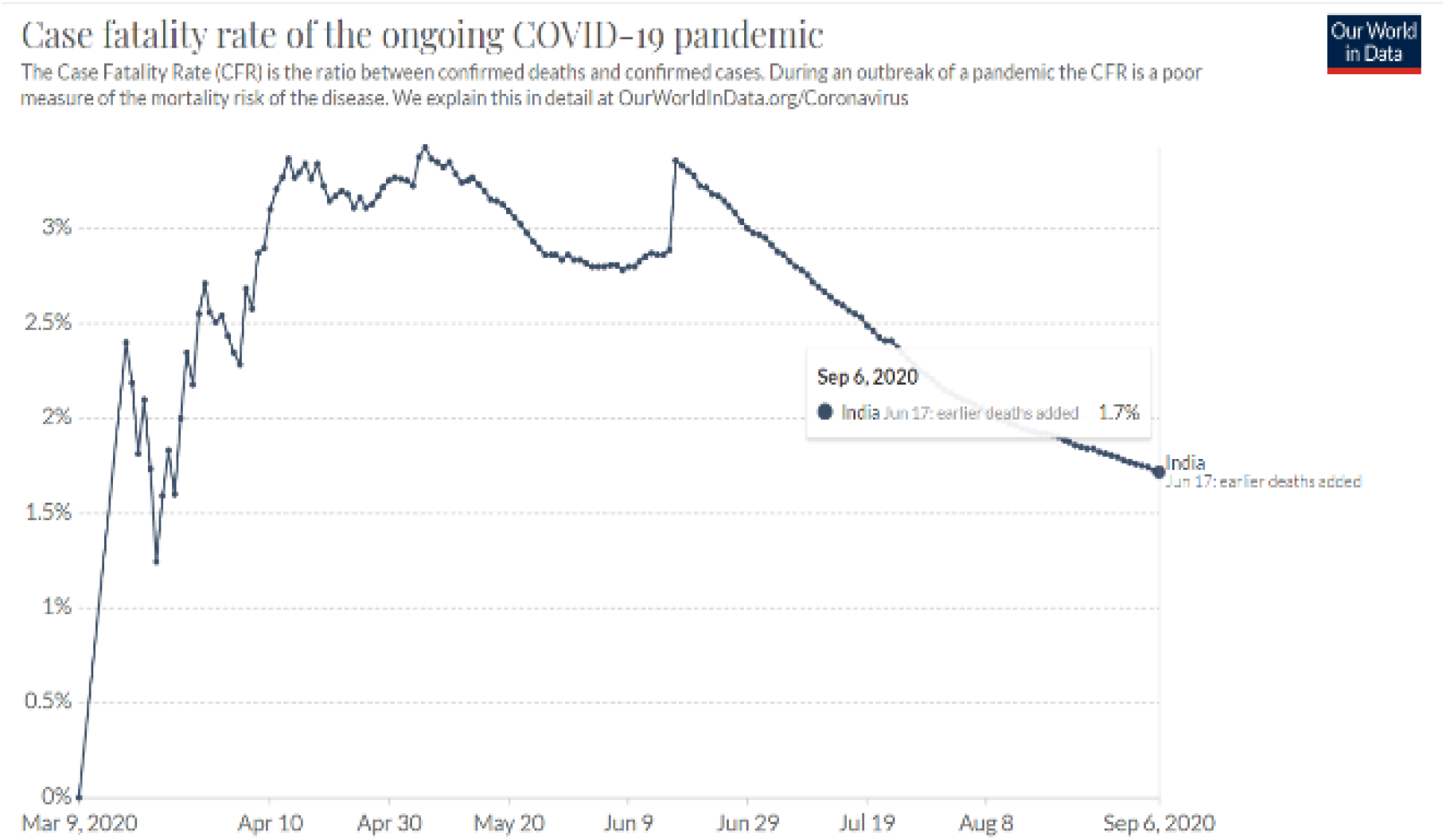
The COVID-19 Case Fatality Rate in India is 1.7%. Source: European Centre for Disease Prevention and Control (Data as on September 6, 2020)^(14)^

Unlike the previous retrospective studies from China, we did not find any significant association between cancer treatment and mortality^(7, 8)^. Our observation is in line with two large cohort studies conducted by Lee *et al*. on 800 COVID-19 positive cancer patients from U K Coronavirus Cancer Monitoring Project, and by Robilotti *et al*. on 423 symptomatic COVID-19 cancer patients at Memorial Sloan Kettering Cancer Centre in New York^(9, 17)^. The recent multi-center study by Pinato *et al*. in the European cancer patients and the Covid-19 and Cancer Consortium (CCC19) database study also strengthens the notion that cancer treatment is not associated with mortality^(10, 18)^. Similar to these studies we found that it is the underlying comorbid conditions and COVID-19 disease severity that are associated with adverse outcomes. Consistent with the CCC19 study we observed a higher burden of preexisting comorbidities (>1) to be associated with increased mortality^(18)^. Cardiovascular diseases and diabetes posed a higher risk of death in our cohort. It should be noted that comorbid conditions like diabetes frequently co-occur with hypertension or coronary artery disease in cancer patients and can further weaken the immune response escalating the risk of death due to COVID-19^(19, 20)^.

## Conclusion

Our study highlights the incredibly high rates of COVID-19 in cancer patients. More distressingly the CFR is 7.6 times more than the national average CFR for COVID-19. Anticancer therapies did have a significant effect on mortality in the cohort. Pre-existing comorbidities specially diabetes and more than one comorbidity are significantly linked with COVID-19 related deaths in cancer patients.

## Data Availability

Data will be made available on a reasonable request to the corresponding authors.

---

Supplementary Table 1.

**Supplementary table 1.**
| **Variable** | **Odds ratio** | **95% CI** | ***P* value** |
| --- | --- | --- | --- |
| Age | 0.99 | 0.96 - 1.02 | 0.40 |
| Comorbidity | 3.16 | 1.16 - 8.61 | 0.02 |
| Chemotherapy, Yes/ No | 1.63 | 0.64 - 4.15 | 0.30 |
| Radiotherapy, Yes/ No | 0.19 | 0.02 - 1.59 | 0.13 |
| Targeted therapy, Yes/ No | 2.70 | 0.42 - 17.37 | 0.29 |
| Immunotherapy, Yes/ No | 0.40 | 0.04 - 3.56 | 0.41 |
| Surgery, Yes/ No | 0.87 | 0.25 - 2.94 | 0.82 |

